# Performance of vaccination with CoronaVac in a cohort of healthcare workers (HCW) - preliminary report

**DOI:** 10.1101/2021.04.12.21255308

**Authors:** Elizabeth de Faria, Ana Rubia Guedes, Maura S. Oliveira, Moacyr Vergara de Godoy Moreira, Fernando Liebhart Maia, Antonio dos Santos Barboza, Mariana Deckers Leme, Leila S. Harima Letaif, Anna Miethke-Morais, Eloisa Bonfá, Aluisio C. Segurado, Francis M. Tomazini, Alcir Alves dos Santos, Pedro Figueiredo, Pâmela dos Santos Andrade, Franciane Mendes de Oliveira, Raissa Heloisa de Araújo Eliodoro, Jaqueline Goes de Jesus, Carolina dos Santos Lazari, Ester C. Sabino, Silvia F. Costa, Antonio Carlos Pedroso de Lima, Anna S. Levin

**Author notes:** **Corresponding author:** Anna S. Levin, Address: Rua Banibas, 618; Sao Paulo-SP; CEP: 05460-010; Brazil.

## Abstract

**Background:** CoronaVac, a vaccine containing inactivated SARS-CoV-2, demonstrated efficacy of 50.39% 14 days or more after the 2nd dose.

The objective of this study is to report the occurrence of symptomatic COVID-19 in a cohort of HCW vaccinated with CoronaVac and to estimate its effectiveness.

**Methods:** CoronaVac was given to HCWs inHospital das Clinicas on 18-21 January, 2021 (epi week 3) (22,402 HCWs), and on 14-16 February, 2021 (epi week 7) (21,652 HCWs). Weekly cases of symptomatic COVID-19 were evaluated. Using the period from 2020 epi week 24 through 2021 epi week 2 (before vaccination), a Poisson regression was fit to model the HCWs with COVID-19 of the hospital, and the officially reported cases in the city of São Paulo. The predicted numbers of cases among HCWs for 2021 epi weeks 3-12 were then compared to the observed numbers of cases (after vaccination). Effectiveness was estimated for weeks 9-12 (2 to 5 weeks after the 2nd dose). 142 samples after vaccination were evaluated for SARS-CoV-2 variants of concern.

**Results:** Since the 1st dose there were 380 HCW diagnosed with COVID-19. On visual analysis, the number of cases of COVID-19 in the city increased sharply in 2021. The number of cases among the HCW did not follow. The estimated effectiveness 2 and 3 weeks after 2nd dose was 50.7% and 51.8%, respectively, and increased over the next 2 weeks. 67/142 samples (47%) were variants of concern, mostly P1 (57).

**Conclusion:** Coronavac is effective in preventing COVID-19.

## Introduction

CoronaVac, a vaccine containing inactivated SARS-CoV-2, was well tolerated and induced humoral responses in phase 1 and 2 studies [1]. Interim analysis of phase 3 clinical trial conducted in Brazil evaluating 12,607 participants demonstrated efficacy of 50.39% (95%CI: 35.26 – 61.98; p=0.005) when considering 15 days or more after the 2nd dose [2].

Efficacy and effectiveness of vaccines measure the proportion of reduction of a disease among vaccinated persons. The difference is that efficacy is used during a clinical trial, under ideal circumstances and effectiveness is used when a study is performed under everyday conditions, outside of clinical trials [3]. To our knowledge there are no studies on the effectiveness of CoronaVac.

The objective of this study is to report the occurrence of symptomatic COVID-19 in a cohort of healthcare workers (HCW) vaccinated with CoronaVac and to estimate its effectiveness.

## Methods

Hospital das Clinicas (HC) is a tertiary-care teaching hospital of the University of São Paulo, Brazil. It is located in the city of São Paulo that has 12,325,232 inhabitants [4]. HC has 2,200 beds distributed over 7 buildings. It was the main reference hospital for severe COVID-19 infections in the State of São Paulo, from 30 March through August 30, 2020, with approximately 1,000 beds dedicated to COVID-19, of which 300 were in intensive care units (ICU). During these months HC admitted approximately 4,500 severe COVID-19 cases. After that period, approximately 25% (555 beds) of HC’s beds, including 200 ICU beds, remained dedicated to COVID-19, while the rest was used for admission of other severe clinical and surgical cases that had backed-up during the previous months. Starting on 2021 epi week 10, the pressing demand led to progressive increases in beds allocated for COVID-19. The work force in HC is approximately 20,000 HCWs who deliver direct assistance or have other close contact with patients.

Since the beginning of the COVID-19 pandemic, symptomatic HCWs were directed to specific HCW health services where they were evaluated by a doctor and received a 3-day leave. On day 3 of symptoms they collected oro-nasopharyngeal swabs, tested by RT-PCR for SARS-CoV-2. If negative and asymptomatic they returned to work. If positive, they received paid leave for a total of 14 days. No follow-up testing was done once the HCW was found to be positive.

Over 4 days (18-21 January, 2021 -epi week 3), a large vaccination effort led to the vaccination of 22,402 HCWs with the first dose of CoronaVac. The second dose was delivered over 3 days (14-16 February, 2021 -during epi week 7) and 21,652 HCWs received the 2nd dose of the vaccine.

CoronaVac is an inactivated SARS-CoV-2 vaccine developed by Sinovac Life Sciences (Beijing, China). Phase 1/2 studies showed it to be safe and to lead to seroconversion [1]. It was licenced for use in Brazil on 17 January, 2021. Unpublished efficacy has been reported to be 50.39% [2]. CoronaVac vaccination was studied using 2 doses of 600 SU administered 2 to 4 weeks apart.

Over the period from 23 February, 2020 (2020 epidemiological week 9) through 28 March, 2021 (2021 epi week 12) we recorded the weekly numbers of HCWs in HC who had symptomatic COVID-19 confirmed by RT-PCR, evaluated by the hospital’s HCW health services.

Over the same period we evaluated the weekly number of COVID-19 cases and COVID-19 related deaths in the city of São Paulo that is publicly reported [5]; and the index of immobility [6]. This index is based on the location of cell phones, with data from the 4 mobile phone companies that cover the city. The cell phones are considered to have “slept” in the location registered between 10PM to 2AM. After that, displacements of more than 200m from that point are registered. The index is the proportion of telephones that were not displaced over 24 hours. We presented the average for each epi week.

RT-PCR was done in samples obtained by swabbing of oro-nasopharynx, using a laboratory developed test based on the Charitè protocol [7]. Part of the positive samples was evaluated for the presence of SARS-CoV-2 variants of concerns (VOC) using a screening protocol [8] that detects the deletion in the NSP6 gene that is common for the P1, B.1.1.7, and B.1.351 lineages. The samples were also studied using a protocol [9] that is able to distinguish B1.1.7 from P1 and B 1.351 lineages.

## Statistical Analysis

A Poisson regression model was used to study the relationship between the weekly number of COVID-19 cases among HC HCW, taken as dependent variable, and cases recorded for the general population of São Paulo city. The fit was based on data from the pre-vaccination period, from 2020 epi week 24 to 2021 epi week 2. Residual analyses were then performed, showing two influential points. After removal of such values, a final model was obtained and used to extrapolate the number of cases among HC HCW considering the known values for the population of São Paulo city in the post-vaccination period (2021 epi week 3 to 12). In addition, 95% prediction intervals were computed using the bootstrap resampling method. The observed values of COVID-19 cases among HC HCW were then compared to the predicted ones for the post-vaccination period. Effectiveness estimates and corresponding 95% bootstrap confidence intervals were computed assuming the observed number of HC HCW cases as the vaccinated group and the predicted numbers from the Poisson model as the unvaccinated group.

## Results

The weekly behaviour of symptomatic COVID-19 among the hospital’s HCW, and the publicly available data on the COVID-19 cases in the city of São Paulo can be seen in Figure 1. The results of deaths in the city of São Paulo and the immobility index, can be seen in Supplemental Figure S1.

**Figure 1:**
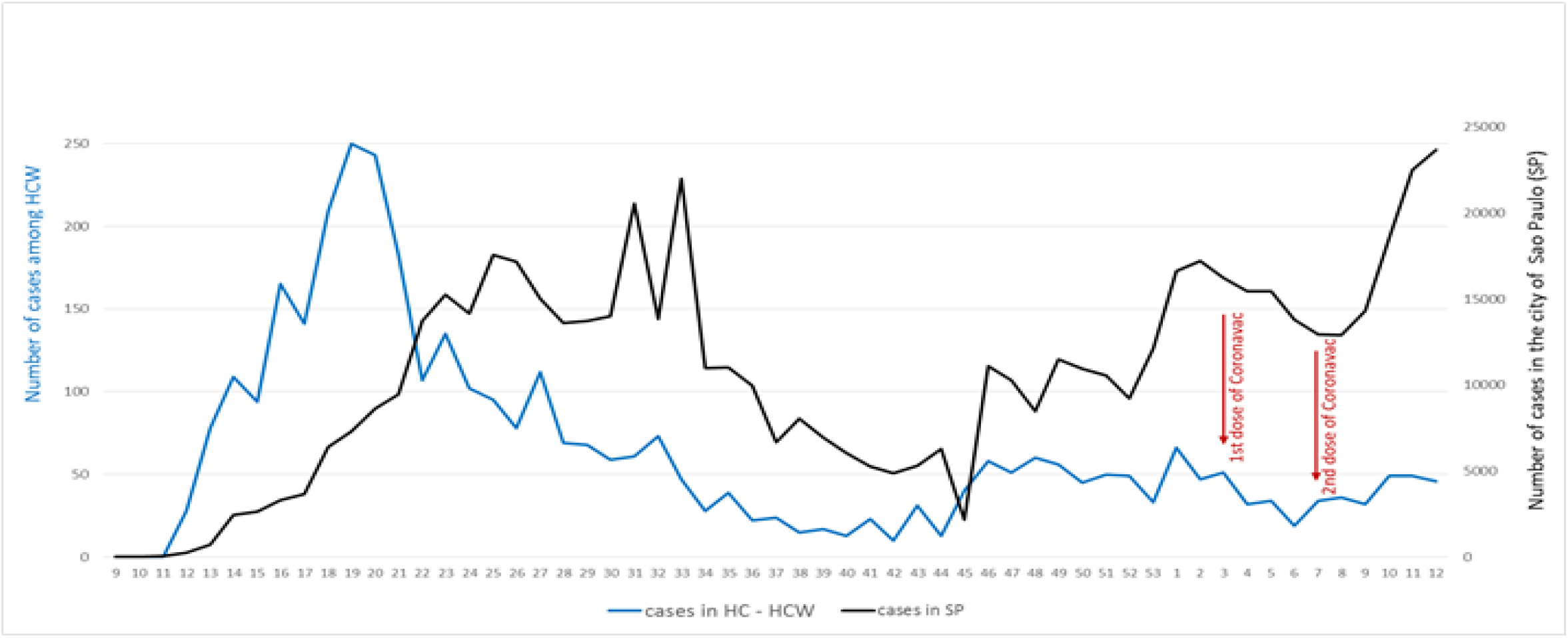
Graphical representation of the number of weekly COVID-19 cases in the city of São Paulo (SP) and among healthcare workers (HCW) of Hospital das Clinicas (HC) (University of São Paulo). Arrows mark the adminstration of the 2 doses of CoronaVac in HC’s HCW.

During the period from 10 March, 2020 through 28 March, 2020 there were 20,187 medical consultations of suspected cases and follow-up of COVID-19 cases at the HCW health facility; 13,713 RT-PCR tests performed on HCW, of which 3,565 (26%) were positive. The 1st peak of cases among HC HCWs preceded the peak of cases in the city. However, the curve of deaths in the city behaved in a similar way to HCW cases. Thus we attributed the difference to sub notification of cases in the city, as testing was not readily available at the beginning of the pandemic. Because of this, in the analysis we excluded 2020 epi weeks 9 through 23. On visual analysis, after HCW vaccination in HC, the number of cases of COVID-19 in the city of São Paulo increased very sharply. The number of cases among the HCW of HC did not follow this pattern.

The results from the Poisson regression fit considering period from 2020 epi week 24 to 2021 epi week 2 can be seen in Figure 2. The observed number of cases for HC HCWs for the weeks 3 through 12 post-vaccination period, as well as prediction values and 95% prediction intervals can be seen in Table 1. The predicted number of cases among HC HCWs can be thought of as what would be observed if this population had not been vaccinated; therefore, the differences between the actual number of cases and the predicted ones may be interpreted as the effect of immunization. None of the prediction intervals includes the observed values, that were always below the interval inferior limit, reinforcing our findings.

**Table 1:** Number of cases of COVID-19 in healthcare workers (HCW) of Hospital das Clinicas (HC), and predicted cases in HCW based on the reported cases in the city of São Paulo, after vaccination with CoronaVac

| Epi week 2021 | Number of cases, city of São Paulo | Number of cases among HC HCWs |  | Prediction interval (95%) |  |
| --- | --- | --- | --- | --- | --- |
|  |  | Observed | Predicted | Inferior limit | Superior limit |
| 3 (1st dose) | 16,232 | 51 | 79.5 | 59.0 | 108.0 |
| 4 | 15,432 | 32 | 73.0 | 55.0 | 94.1 |
| 5 | 15,460 | 34 | 73.2 | 53.0 | 97.0 |
| 6 | 13,788 | 19 | 61.2 | 45.0 | 80.0 |
| 7 (2nd dose) | 12,954 | 34 | 56.0 | 42.0 | 72.0 |
| 8 | 12,906 | 36 | 55.7 | 40.0 | 74.0 |
| 9 | 14,332 | 30 | 64.9 | 47.0 | 85.0 |
| 10 | 18,536 | 49 | 101.6 | 71.0 | 147.0 |
| 11 | 22,511 | 49 | 155.2 | 102.0 | 267.1 |
| 12 | 23,889 | 46 | 175.6 | 110.0 | 317.1 |

**Figure 2:**
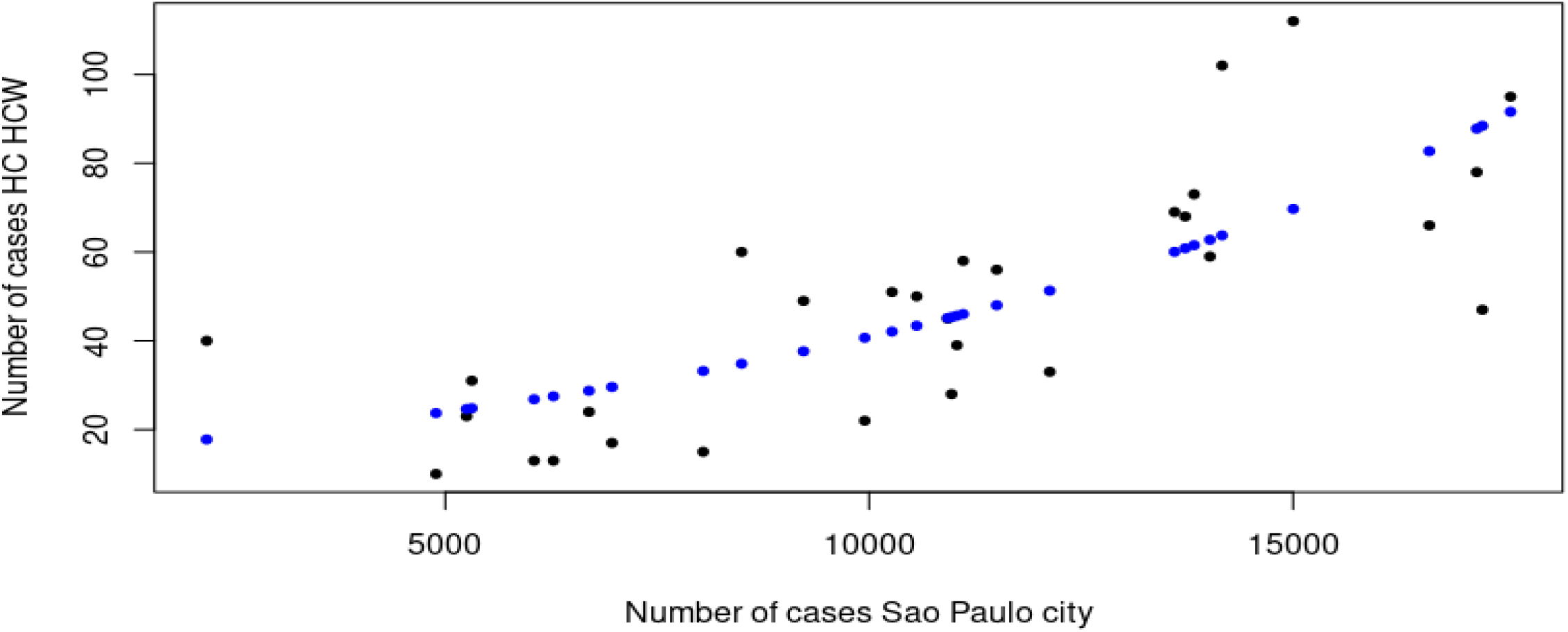
Scatterplot for observed (black) and predicted values, based on the Poisson regression model (blue), for weekly cases of COVID-19 among healthcare workers of Hospital das Clinicas (HC HCW) (2020 epidemiological week 24 to 2021 epi week 2). Axis X refers to the weekly number of cases in the city of Sao Paulo, Brazil.

Estimated effectiveness was 50.7% (95% CI: 33.3-62.5%); 51.8% (95%CI: 30.0-66.0%); 68.4% (95%CI: 51.0-80.8%); and 73.8% (95%CI: 57.0-84.8%) for 2021 epi weeks 9, 10, 11, and 12, respectively. These weeks correspond to 2, 3, 4, and 5 weeks after the 2nd dose of CoronaVac, respectively. The estimated effectiveness on epi weeks 9 and 10 was somewhat similar to the efficacy described for this vaccine; and for weeks 11 and 12 it was even greater. Nevertheless, estimated effectiveness for weeks 10-12 should be interpreted with caution because the number of cases in the city of São Paulo was superior to the maximum number of cases (17,566) considered in estimating the Poisson regression used for predictions.

During 2021, there were 9 hospitalizations of HCW in HC due to COVID-19: 6 had not been vaccinated, 1 had received one dose of CoronaVac, and 2 had received 2 doses. Only one of these died, who had not been vaccinated.

Among 142 HCW’s respiratory samples randomly evaluated, in 67 (47%) variants of concern (VOC) were detected: 57 were variant P1, 5 were B.1.1.7, and 5 were other VOC not identified by our methods.

The index of immobility increased at the beginning of the pandemic reaching a peak of 57%. Slowly it decreased and has been relatively stable since November 2020, at about 40% (Supplemental Figure S1).

By 29 March, 2021, 1,255,627 people (10% of the population) in the city of São Paulo had received at least one dose of a COVID-19 vaccine, and 437,438 (3.5%) had received 2 doses [10].

## Discussion

Despite the studies of efficacy of vaccines against COVID-19, little is known about their effectiveness, or how they perform in real-life conditions. In a large cohort of HCWs who were vaccinated in a mass campaign with CoronaVac, an inactivated SARS-CoV-2 vaccine, we observed an important decrease over time of confirmed symptomatic cases of COVID-19 in relation to what would have been expected considering the epidemiological situation in the same community.

The phase 3 trial evaluating CoronaVac included over 12,000 individuals and showed a 50.38% efficacy [2]. The data are still unpublished.

Following licencing in Brazil, mass vaccination of HC HCWs was done. This gave us an opportunity to estimate the effectiveness of the vaccine in a large cohort. The preliminary results suggest that it was effective.

Although efficacy has been studied and varied among the different vaccines, little has been published on effectiveness of COVID-19 vaccines, that is their effect in real-world conditions. An evaluation of 2 FDA approved vaccines (BNT162b2 from Pfizer/BioNTech and mRNA-1273 from Moderna, with efficacies of 95.0% and 94.1%, respectively) in the Mayo Clinic system in the US, showed that the administration of 2 doses was 88.7% effective in preventing SARS-CoV-2 infection at least 36 days after the 1st dose [11]. A study involving 6 locations in the US, with HCW and first responders, showed an effectiveness of 90% after 2 doses of mRNA vaccines [12]. A study in Israel, a country that has been highly successful in rapidly vaccinating its population with BNT162b2, estimated an effectiveness of 66-85% in reducing SARS-CoV-2 positive cases and over 90% in reducing severe hospitalizations [13]. To our knowledge, there has been no data on the effectiveness of CoronaVac, an inactivated SARS-CoV-2 vaccine. The estimated effectiveness of CoronaVac in our study was at least 50% starting 2 weeks after the second dose.

Another interesting finding was the prevalence of variants: almost half of the studied samples. There has been a concern whether vaccination can lead to an increase in infections caused by variants because vaccines may not be efficient against them [14]. However, we believe that this high prevalence, especially of P1, reflects the epidemiological situation in the region. The variant P1 was described as emerging in the Amazon region in Brazil in November 2020 and spread rapidly, becoming the predominant strain in 7 weeks, accounting for 87% of cases [15]. It has also dominated the epidemic in several regions of the country. Recently the city of São Paulo reported 64% of P1 and 7% of B.1.1.7 in samples collected from 16 February to 6 March, 2021 [16]. Thus, our results probably reflect the distribution of variants in the community.

Our study has limitations. In the first place, it is a very preliminary evaluation in which details of affected patients such as severity of disease and deaths were not taken into account. Secondly, the peak of cases among HCWs in HC preceded the peak of cases in the city of São Paulo. We believe that this was due to the lack of diagnosis of mild and moderate cases in the city. In HC, testing was immediately available to all symptomatic HCWs since the beginning of the pandemic. In support of our idea, the peak of deaths in the city was similar to the cases in our HCWs. It has been discussed that deaths are a more reliable surveillance tool in situations in which testing capacity is limited [17, 18] which was especially true in the beginning of the pandemic in São Paulo. Considering this situation we chose not to include data from 2020 epi weeks 9 through 23 when designing our predictive model. As mentioned earlier, the results for 2021 epi weeks 10-12 should be interpreted with caution, as the numbers of cases in the city of São Paulo in these weeks were outside the range variation observed during parameter estimation of the Poisson regression model.

In conclusion, since 2021 epi week 3, when the first dose of CoronaVac was administered to the HCW of the hospital, COVID-19 cases among our HCWs were lower than expected. São Paulo started a second wave of COVID-19 soon after the completion of vaccination in HC, that was not reflected among our HCWs.

## Data Availability

Data is available upon request.

## References

1- Wu Z, Hu Y, Xu M, Chen Z, Yang W, Jiang Z, Li M, Jin H, Cui G, Chen P, Wang L, Zhao G, Ding Y, Zhao Y, Yin W.Safety, tolerability, and immunogenicity of an inactivated SARS-CoV-2 vaccine (CoronaVac) in healthy adults aged 60 years and older: a randomised, double-blind, placebo-controlled, phase 1/2 clinical trial. Lancet Infect Dis On-line: February 3, 2021. https://doi.org/10.1016/S1473-3099(20)30987-7.

2- Agência Nacional de Vigilância Sanitária (Brazilian agency for drug approval). Autorização de Uso Emergencial de Vacinas contra a COVID-19. Available at: https://www.gov.br/anvisa/pt-br/assuntos/noticias-anvisa/2021/confira-materiais-da-reuniao-extraordinaria-da-dicol/1-apresentacao-ggmed-coronavac.pdf

3- Singal AG, Higgins PDR, Waljee AK. A Primer on Effectiveness and Efficacy Trials. Clinical and Translational Gastroenterology 2014; 5: e45. doi:10.1038/ctg.2013.13

4- Instituto Brasileiro de Geografia e Estatística. Cidades e Estados. Available at: https://www.ibge.gov.br/cidades-e-estados/sp/sao-paulo.html).

5- State of São Paulo, Brazil. Dados abertos. Registro de casos e óbitos por município e data de notificação no Estado de São Paulo. Available at: https://www.saopaulo.sp.gov.br/planosp/simi/dados-abertos/

6- State of São Paulo, Brazil. Adesão ao isolamento social em SP. Available at: https://www.saopaulo.sp.gov.br/coronavirus/isolamento/

7- Corman VM, Landt O, Kaiser M, Molenkamp R, Meijer A, Chu DK, Bleicker T, et al. Detection of 2019 novel coronavirus (2019- nCoV) by real-time RT-PCR Euro Surveill (3) (2020), Article 2000045

8- Romano CM, Jesus J, Felix AC, Paula AV, Andrade PS, Oliveira FM, Cândido D, Faria N, Souza WM, Sabino EC. Real-Time PCR protocol to screen for SARS-COV-2 variants of concern (B.1.1.7, P.1 and B.1.1.35). https://www.protocols.io/view/real-time-pcr-protocol-to-screen-for-sars-cov-2-va-bszbnf2n?step=8

9- Vogels CBF, Breban MI, Alpert T, Petrone ME, Watkins AE, Ott IM, Jesus JG, Claro IM, et al. PCR assay to enhance global surveillance for SARS-CoV-2 variants of concern. medRxiv 2021 Mar 12;2021.01.28.21250486. doi:10.1101/2021.01.28.21250486.

10- Cidade de São Paulo. Boletim Diário COID-19 (no 368). 29 March, 2021. Available at: https://www.prefeitura.sp.gov.br/cidade/secretarias/upload/saude/20210329_boletim_covid19_diario.pdf

11- Pawlowski C, Lenehan P, Puranik A, Argawal V, Venkatakrishnan AJ, Niesen MJM, O’Horo JC, Badley AD, Halamka J, Soundararajan V. FDA-authorized COVID-19 vaccines are effective per real-world evidence synthesized across a multi-state health system. medRxiv preprint doi:https://doi.org/10.1101/2021.02.15.21251623

12- Thompson MG, Burgess JL, Naleway AL, Tyner HL, Yoon SK, Meece J, Olsho LEW, et al. Interim Estimates of Vaccine Effectiveness of BNT162b2 and mRNA-1273 COVID-19 Vaccines in Preventing SARS-CoV-2 Infection Among Health Care Personnel, First Responders, and Other Essential and Frontline Workers — Eight U.S. Locations, December 2020–March 2021. Morbidity and Mortality Weekly Report vol 20, Early release. March 29, 2021. Available at: https://www.cdc.gov/mmwr/volumes/70/wr/mm7013e3.htm

13- Aran D. Estimating real-world COVID-19 vaccine effectiveness in Israel. medRxiv preprint doi:https://doi.org/10.1101/2021.02.05.21251139

14- World Health Organization. Science in 5. Episode #31-Vaccines, variants & doses (Katherine O’Brien, Vismita Gupta-Smith). Available at: https://www.who.int/emergencies/diseases/novel-coronavirus-2019/media-resources/science-in-5/episode-31---vaccines-variants-and-doses

15- Faria NF, Mellan TA, Whittaker C, Claro IM, Candido DS, Mishra S, Crispim MAE, Sales FC, et al. Genomics and epidemiology of a novel SARS-CoV-2 lineage in Manaus, Brazil. Preprint doi:10.1101/2021.02.26.21252554

16- Cidade de São Paulo. Monitoramento de novas variantes de SARS-CoV-2 no Município de São Paulo. 26 March, 2021. Available at: https://www.prefeitura.sp.gov.br/cidade/secretarias/upload/saude/situacao_covid19_03_26_03_2021.pdf

17- Mwananyanda L, Gill CJ, MacLeod W, Kwenda G, Pieciak R, Mupila Z, Lapidot R, Mupeta F, Forman L, Ziko L, Etter L, Thea D. Covid-19 deaths in Africa: prospective systematic postmortem surveillance study. BMJ 2021;372:334. doi:10.1136/bmj.n334

18- Freitas ARR, Medeiros NM, Frutuoso LCV, Beckedorff OA, Martin LMA, Coelho MMM, Freitas GGS, Lemos DRQ, Cavalcanti LPG. Tracking excess deaths associated with the COVID-19 epidemic as an epidemiological surveillance strategy-preliminary results of the evaluation of six Brazilian capitals. Rev Soc Bras Med Trop 2020; 53: e20200558. doi:10.1590/0037-8682-0558-2020

