## Supplemental Figure S1 for "Performance of vaccination with CoronaVac in a cohort of healthcare workers (HCW) - preliminary report"

**Supplemental Figure S1**: Weekly number of COVID-19 related deaths, and weekly index of immobility - City of Sao Paulo, Brazil (2020 epidemiological week 9 - 2021 epidemiological week 12)

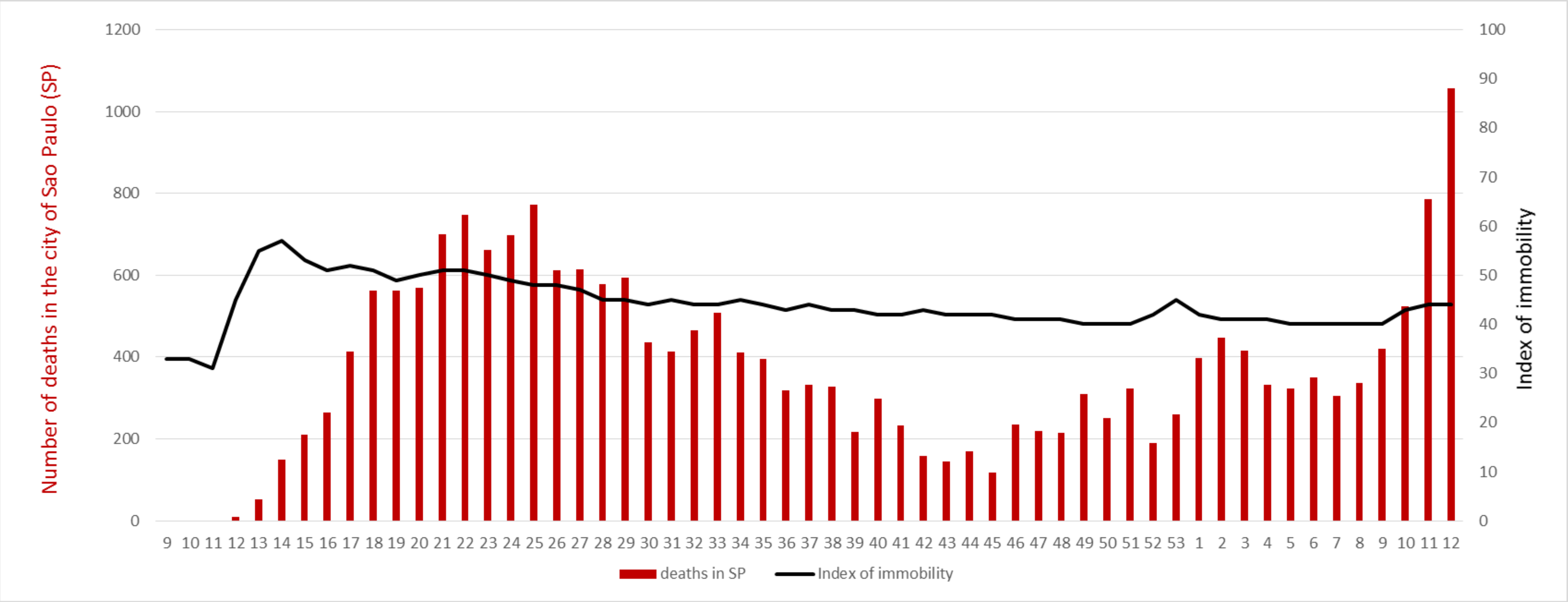
